# Inherited basis of visceral, abdominal subcutaneous and gluteofemoral fat depots

**DOI:** 10.1101/2021.08.24.21262564

**Authors:** Saaket Agrawal, Minxian Wang, Marcus D. R. Klarqvist, Joseph Shin, Hesam Dashti, Nathaniel Diamant, Seung Hoan Choi, Sean J. Jurgens, Patrick T. Ellinor, Anthony Philippakis, Kenney Ng, Melina Claussnitzer, Puneet Batra, Amit V. Khera

## Abstract

For any given level of overall adiposity – as commonly quantified by body mass index (BMI) within clinical practice – individuals vary considerably in fat distribution. We and others have noted that increased visceral fat (VAT) is associated with increased cardiometabolic risk, while gluteofemoral fat (GFAT) may be protective. Familial partial lipodystrophy (FPLD) – often caused by rare variants in the *LMNA* gene – represents an extreme example of this paradigm, leading to a severe shift to visceral fat with subsequent insulin resistance and adverse metabolic profile. By contrast, the inherited basis of body fat distribution in the broader population is not fully understood. Here, we studied up to 38,965 UK Biobank participants with VAT, abdominal subcutaneous (ASAT), and GFAT volumes precisely quantified using abdominal MRI. Because genetic associations with these raw depot volumes were largely driven by variants known to affect BMI, we next studied six phenotypes of local adiposity: VAT adjusted for BMI (VATadjBMI), ASATadjBMI, GFATadjBMI, VAT/ASAT, VAT/GFAT, and ASAT/GFAT. We identify 178 unique loci associated with at least one adiposity trait, including 29 newly-identified loci. Rare variant association studies extend prior evidence of association for *PDE3B* as an important modulator of fat distribution. Sex-specific analyses of local adiposity traits noted overall higher estimated heritability in females, increased effect sizes for identified loci, and 25 female-specific associations. Individuals in the extreme tails of fat distribution phenotypes were highly enriched for predisposing common variants, as quantified using polygenic scores. Taking GFATadjBMI as an example, individuals with extreme values were 3.8-fold (95%CI 2.8 to 5.2) more likely to have a polygenic score within the top 5% of the distribution. These results – using more precise and BMI-independent measures of local adiposity – confirm fat distribution as a highly heritable trait with important implications for cardiometabolic health outcomes.

## INTRODUCTION

Overall fat mass and fat distribution represent two distinct axes of variation that determine the health impacts of adipose tissue. Individuals with higher body mass index (BMI) – defining obesity – are at elevated risk of type 2 diabetes and cardiovascular events, but increased cardiometabolic risk has also been noted in individuals with the same BMI when fat is disproportionally depleted in more favorable gluteofemoral fat depots and deposited instead in visceral and ectopic fat depots.^1–5^ An extreme example of this paradigm occurs in Mendelian lipodystrophies, such as those caused by missense mutations in the *LMNA* and *PPARG* genes.^6–10^ By contrast, the genetic architecture of more subtle variation in fat distribution across the general population warrants further attention.

In general, prior studies aiming to elucidate common genetic variation contributing to fat distribution can be categorized into three study types: (1) common-variant association studies (CVAS) on anthropometric proxies of fat distribution, (2) studies combining CVAS summary statistics of metabolic and anthropometric traits, and (3) CVASs on imaging-based measures of fat distribution. The first type has been spearheaded by the GIANT consortium and others, which identified over 300 loci associated with waist-to-hip ratio adjusted for BMI (WHRadjBMI) in an analysis of nearly 700,000 individuals.^11, 12^ Another recent CVAS aimed to examine fat distribution using estimates of body composition based on stepping on a scale equipped with impedance technology, known to be reasonably accurate for total fat volume but less so for fat distribution.^13–15^ Despite the considerable value of these studies, a central limitation is an unclear relationship between each anthropometric trait and each fat depot of biological interest – for example, an increase in WHRadjBMI could be capturing an increase in visceral adipose tissue (VAT; around the abdominal organs), an increase in abdominal subcutaneous adipose tissue (ASAT; belly fat under the skin), or a decrease in gluteofemoral adipose tissue (GFAT; hip and thigh fat), or some combination of these perturbations.^16, 17^

A second category of studies has aimed to gain further resolution into anthropometric loci by combining summary statistics of metabolic and anthropometric traits, generating clusters of metabolically favorable and unfavorable loci.^18–22^ These studies have succeeded in establishing a common variant basis for metabolically distinct fat depots, with seminal work demonstrating that an insulin resistance polygenic score is associated with lower hip circumference in the general population, and that individuals with familial partial lipodystrophy type 1 (FPLD1) have a higher burden of this polygenic score.^19^ Along with their reliance on anthropometric proxies of fat distribution, these studies are limited by their inclusion requirement of nominal significance across multiple metabolic traits which is likely leading to only a fraction of the genetic architecture of fat distribution being described.

Finally, the third category of studies performed CVASs on imaging-derived measurements of fat depots. These include CVASs of CT-quantified VAT and ASAT in nearly 20,000 individuals, CVASs on MRI-quantified VAT and ASAT, and a CVAS of a predicted VAT trait using several anthropometric traits trained on over 4,000 DEXA-measured VAT values.^23–26^ These studies have been important for translating insights from anthropometric and metabolic trait CVASs to image-derived measurements of the fat depots of interest, but have been limited by (1) the absence of GFAT, which appears to have a metabolically protective role in contrast to VAT and ASAT, and frequently (2) a reliance on raw, unadjusted fat depot metrics which are highly correlated with both each other and BMI.

In this study, we investigate the common and rare variant genetic architecture of local fat depots as quantified by MRI in up to 38,965 UK Biobank participants. Beyond study of raw VAT, ASAT, and GFAT volumes, we analyze six traits that better reflect local adiposity and fat distribution: VATadjBMI, ASATadjBMI, GFATadjBMI, VAT/ASAT, VAT/GFAT, and ASAT/GFAT. This study is the largest imaging-based study to date to disentangle the genetic architecture of different fat depots using these local adiposity metrics and the first CVAS to date of GFAT, a fat depot that appears to confer *protection* from adverse cardiometabolic health.^5, 27^

## RESULTS

Visceral adipose tissue (VAT), abdominal subcutaneous adipose tissue (ASAT), and gluteofemoral adipose tissue (GFAT) volumes were quantified in participants of the UK Biobank using a deep learning model trained on two-dimensional projections of body MRIs, as previously described.^5^ Among those with MRI-quantified fat depot volumes, 39,076 had genotyping array data available, enabling common variant association studies in up to 38,965 participants after quality control (**Supplementary Figure S1**). Mean age in the genotyped cohort was 64.5 years, 51% were female, and 97% were white (**Supplementary Table S1**). As expected, significant sex differences in fat depot volumes were observed – males had higher mean VAT volume (5.0 L versus 2.6 L), while females had higher ASAT volume (7.9 versus 5.9 L) and GFAT volume (11.3 versus 9.3 L).^28, 29^

Six additional adiposity traits – designed to better capture local adiposity and fat distribution – were additoinally computed for each individual: VATadjBMI, ASATadjBMI, GFATadjBMI were computed by taking sex-specific residuals against age, age squared, body mass index (BMI), and height, while VAT/ASAT, VAT/GFAT, and ASAT/GFAT were computed by taking ratios between each pair of fat depots (**Supplementary Figure S2**). While VAT, ASAT, and GFAT volumes were highly correlated with BMI (Pearson *r* ranging from 0.77-0.88), the three BMI-adjusted traits (Pearson r = 0) and three fat depot ratios were only modestly correlated with BMI (Pearson *r* ranging from 0.18-0.56) (**Supplementary Figure S3A-B**), providing useful BMI-independent metrics for downstream analyses.

### Local adiposity traits are highly heritable and genetically distinct from each other

To quantify the inherited component to each of these adiposity traits, we used the BOLT-LMM algorithm to estimate SNP-heritability for each of the 9 adiposity traits. Heritability estimates for VAT, ASAT, and GFAT ranged from 0.30-0.35, comparable to that observed for BMI in the same individuals (h_g_^2^: 0.31) (**Supplementary Table S2**). BMI-adjusted fat depots and fat depot ratios tended to have higher heritability compared to unadjusted fat depots and BMI (h_g_^2^ ranging from 0.34-0.41). In contrast, waist-to-hip ratio adjusted for BMI (WHRadjBMI), an anthropometric proxy for local adiposity, was less heritable than these traits (h_g_ : 0.20). In sex-stratified analyses, most adiposity traits were more heritable in female as compared to male participants, with the greatest heritability across all analyses for GFATadjBMI in females (h_g_ : 0.53).

To study the genetic correlations (r_g_) between the adiposity and related anthropometric traits, we used LD-score regression.^30, 31^ Consistent with observational Pearson correlations, the raw VAT, ASAT, and GFAT volumes were highly genetically correlated with BMI (r_g_ ranging from 0.66-0.82), while the three adjusted-for-BMI fat depots, VAT/ASAT, and VAT/GFAT exhibited low genetic correlation with BMI (r_g_ ranging from -0.16-0.28) (**Supplementary Figure S4A-B**). In sex-combined analyses, VATadjBMI, ASATadjBMI, and GFATadjBMI were genetically correlated with their unadjusted counterparts (r_g_ ranging from 0.45-0.59), but nearly independent of one another (r_g_ ranging from -0.24-0.15), suggesting that adjusted-for-BMI traits can enable fat depot-specific genetic analyses. Finally, WHRadjBMI exhibited positive genetic correlations with VATadjBMI (r_g_: 0.65) and ASATadjBMI (r_g_: 0.25), and a negative genetic correlation with GFATadjBMI (r_g_: -0.29), consistent with the perturbations needed in each fat depot to drive a change in WHRadjBMI.

### Common variant architecture of adiposity traits

We next conducted CVAS for each of the nine adiposity traits – VAT, ASAT, GFAT, VATadjBMI, ASATadjBMI, GFATadjBMI, VAT/ASAT, VAT/GFAT, and ASAT/GFAT – in sex-combined and sex-stratified groups using BOLT-LMM. After genotyping quality control, we tested associations in up to 38,965 participants with 11.5 million imputed SNPs with minor allele frequency (MAF) > 0.005. Across all 27 association studies, 386 locus-trait associations were genome-wide significant at the P-value threshold of 5 x 10^-8^ – 213 of these associations remained significant at a Bonferroni-corrected threshold of 5 x 10^-8^ / 27 = 1.9 x 10^-9^ (**Supplementary Table S4**). When loci across multiple traits in high LD were collapsed into a single locus (R^2^ > 0.7), 178 loci remained, 29 of which were newly-identified (defined as R^2^ < 0.1 with all genome-wide significant associations with prior adiposity and relevant anthropometric traits in the GWAS catalog) (**Table 1**; **Methods**; **Supplementary Tables S5-6**).^32^ Consistent with heritability estimates, the greatest number of loci were identified in association with GFATadjBMI (54 lead SNPs), while the fewest were identified in association with ASAT (6 lead SNPs). The greatest genomic inflation parameter (λ_GC_) was observed with GFATadjBMI (λ_GC_: 1.14) – the LD-score regression intercept was 1.05, consistent with polygenicity rather than significant population structure.^30^

**TABLE 1.** 29 newly-identified loci discovered in this study.

| Trait | CHR | BP | SNP | Effect Allele | Other Allele | EAF | BETA | SE | P-value | Nearest Gene |
| --- | --- | --- | --- | --- | --- | --- | --- | --- | --- | --- |
| GFAT | 11 | 95840436 | rs1074742 | A | G | 0.401 | 0.041 | 0.007 | 1.40E-08 | MAML2 |
| GFAT | 12 | 124344710 | rs138756410 | T | C | 0.986 | -0.172 | 0.031 | 3.00E-08 | DNAH10 |
| GFAT | 12 | 125092343 | rs4765159 | A | G | 0.018 | 0.146 | 0.027 | 3.50E-08 | NCOR2 |
| VATadjBMI | 2 | 121310704 | rs35932591 | C | T | 0.879 | 0.061 | 0.011 | 3.80E-08 | LINC01101 |
| VATadjBMI | 10 | 25767521 | rs1329254 | C | T | 0.37 | 0.042 | 0.007 | 1.40E-08 | GPR158 |
| VATadjBMI | 11 | 69195097 | rs7933253 | T | C | 0.048 | 0.098 | 0.017 | 1.30E-08 | LOC102724265 |
| VATadjBMI<br>(Male) | 2 | 121310704 | rs35932591 | C | T | 0.88 | 0.086 | 0.016 | 3.90E-08 | LINC01101 |
| VATadjBMI<br>(Female) | 3 | 56901687 | rs1500714 | C | G | 0.854 | 0.081 | 0.015 | 1.80E-08 | ARHGEF3 |
| ASATadjBMI | 1 | 201016296 | rs3850625 | G | A | 0.882 | -0.079 | 0.011 | 1.80E-12 | CACNA1S |
| ASATadjBMI | 9 | 1052722 | rs6474550 | G | T | 0.66 | 0.045 | 0.008 | 1.30E-09 | DMRT2 |
| ASATadjBMI | 15 | 62757857 | rs17205757 | A | G | 0.674 | -0.042 | 0.008 | 3.20E-08 | MIR6085 |
| ASATadjBMI | 17 | 76324751 | rs4444401 | A | G | 0.473 | -0.04 | 0.007 | 4.20E-08 | SOCS3 |
| ASATadjBMI<br>(Female) | 1 | 116916645 | rs749166380 | CT | C | 0.102 | 0.102 | 0.018 | 2.20E-08 | ATP1A1 |
| ASATadjBMI<br>(Female) | 8 | 58352327 | rs776481989 | ATAAT | A | 0.998 | 0.795 | 0.134 | 8.60E-09 | LOC101929488 |
| GFATadjBMI | 2 | 226768344 | 2:226768344_CA_C | CA | C | 0.193 | -0.051 | 0.009 | 2.60E-08 | NYAP2 |
| GFATadjBMI | 3 | 196818853 | rs13099700 | A | G | 0.722 | 0.047 | 0.008 | 7.90E-09 | DLG1 |
| GFATadjBMI | 5 | 38810354 | rs142369482 | G | GT | 0.656 | -0.044 | 0.008 | 9.10E-09 | OSMR-AS1 |
| GFATadjBMI<br>(Male) | 4 | 104780790 | rs528845403 | A | AATGT<br>GT | 0.991 | -0.325 | 0.061 | 2.40E-08 | TACR3 |
| GFATadjBMI<br>(Female) | 1 | 181161153 | rs7550430 | A | G | 0.998 | 0.892 | 0.144 | 1.80E-09 | LINC01732 |
| GFATadjBMI<br>(Female) | 2 | 165533198 | rs386652275 | T | TC | 0.974 | -0.19 | 0.034 | 3.20E-08 | COBLL1 |
| VAT/ASAT | 8 | 25459001 | rs3890765 | C | A | 0.941 | -0.084 | 0.015 | 6.80E-09 | CDCA2 |
| VAT/ASAT | 9 | 1054362 | rs6474552 | G | C | 0.432 | -0.04 | 0.007 | 1.20E-08 | DMRT2 |
| VAT/ASAT | 10 | 122992475 | rs11199845 | C | T | 0.46 | 0.055 | 0.007 | 1.50E-14 | FGFR2 |
| VAT/ASAT<br>(Male) | 2 | 61760756 | rs13390751 | A | C | 0.838 | 0.076 | 0.013 | 1.30E-08 | XPO1 |
| VAT/ASAT<br>(Male) | 10 | 122992442 | rs11199844 | C | T | 0.463 | 0.059 | 0.01 | 5.90E-09 | FGFR2 |
| VAT/ASAT<br>(Female) | 12 | 121319417 | rs59757908 | T | C | 0.995 | -0.425 | 0.076 | 4.20E-08 | SPPL3 |
| VAT/GFAT<br>(Female) | 1 | 162430821 | rs9660318 | G | C | 0.203 | 0.068 | 0.012 | 1.80E-08 | UHMK1 |
| VAT/GFAT<br>(Female) | 2 | 116072770 | rs11399916 | T | TA | 0.256 | 0.06 | 0.011 | 3.70E-08 | DPP10 |
| VAT/GFAT<br>(Female) | 6 | 32975699 | rs9276981 | G | C | 0.809 | -0.064 | 0.012 | 4.60E-08 | HLA-DOA |
| ASAT/GFAT | 5 | 55830865 | rs39837 | C | T | 0.667 | 0.043 | 0.007 | 2.60E-08 | LINC01948 |
| ASAT/GFAT | 14 | 95219657 | rs8006225 | G | T | 0.817 | 0.055 | 0.009 | 2.60E-09 | GSC |
| ASAT/GFAT<br>(Female) | 5 | 55830865 | rs39837 | C | T | 0.666 | 0.061 | 0.01 | 9.10E-09 | LINC01948 |

Newly-identified loci were defined as loci that were not in LD ( $R^2 < 0.10$ ) with any of the loci in the GWAS catalog for adiposity or related anthropometric traits (see **Methods**).<sup>32</sup> Note that 32 entries are present rather than 29 because (1) loci could be associated with multiple traits (as is the case with rs35932591 and rs39837) and (2) loci in LD ( $R^2 > 0.7$ ) are reported separately in this table (as is the case with rs11199844 and rs11199845).

We began by investigating the genetic architecture of VAT, ASAT, and GFAT volumes (**Supplementary Figure S5**). All three traits shared a genome-wide significant association with an intronic *FTO* variant (rs56094641) previously associated with childhood and adult obesity.^33–35^ ASAT harbored the strongest association with this locus (P = 1.3 x 10^-22^), followed by GFAT (P = 1.2 x 10^-12^), and finally VAT (P = 3.3 x 10^-10^), reflecting the strength of observational and genetic correlation of each fat depot with BMI. Given evidence from observational and genetic analyses indicating that a large component of each fat depot volume trait is accounted for by BMI – or “overall adiposity” – we limited further common variant analyses to the three adjusted-for-BMI traits and three fat depot ratios, aiming to study the genetic architecture of “local adiposity”.

For VATadjBMI, 30 genome-wide significant associations were identified (P < 5 x 10^-8^) (**Figure 1**; **Supplementary Figure S6**). The two most strongly associated variants were an intronic *CDCA2* variant (rs11992444; P = 1.3 x 10^-29^) previously associated with WHRadjBMI and serum triglycerides, and an intronic *PEPD* variant (rs10406327; P = 3.3 x 10^-24^) previously associated with waist circumference adjusted for BMI (WCadjBMI) and type 2 diabetes.^12, 36–38^ Newly-identified loci in association with VATadjBMI included an intronic *GPR158* variant (rs1329254; P = 1.4 x 10^-8^), and an intronic *ARHGEF3* variant exclusively in females (rs1500714; P = 1.8 x 10^-8^). Prior work has similarly noted female-specific effects of variation in this gene including an association with postmenopausal osteoporosis in humans and ARHGEF3-KO mice being found to have improved muscle regeneration following injury, with an enhanced rate in females, although the role of this gene on fat distribution is uncertain.^39, 40^

**FIGURE 1.**
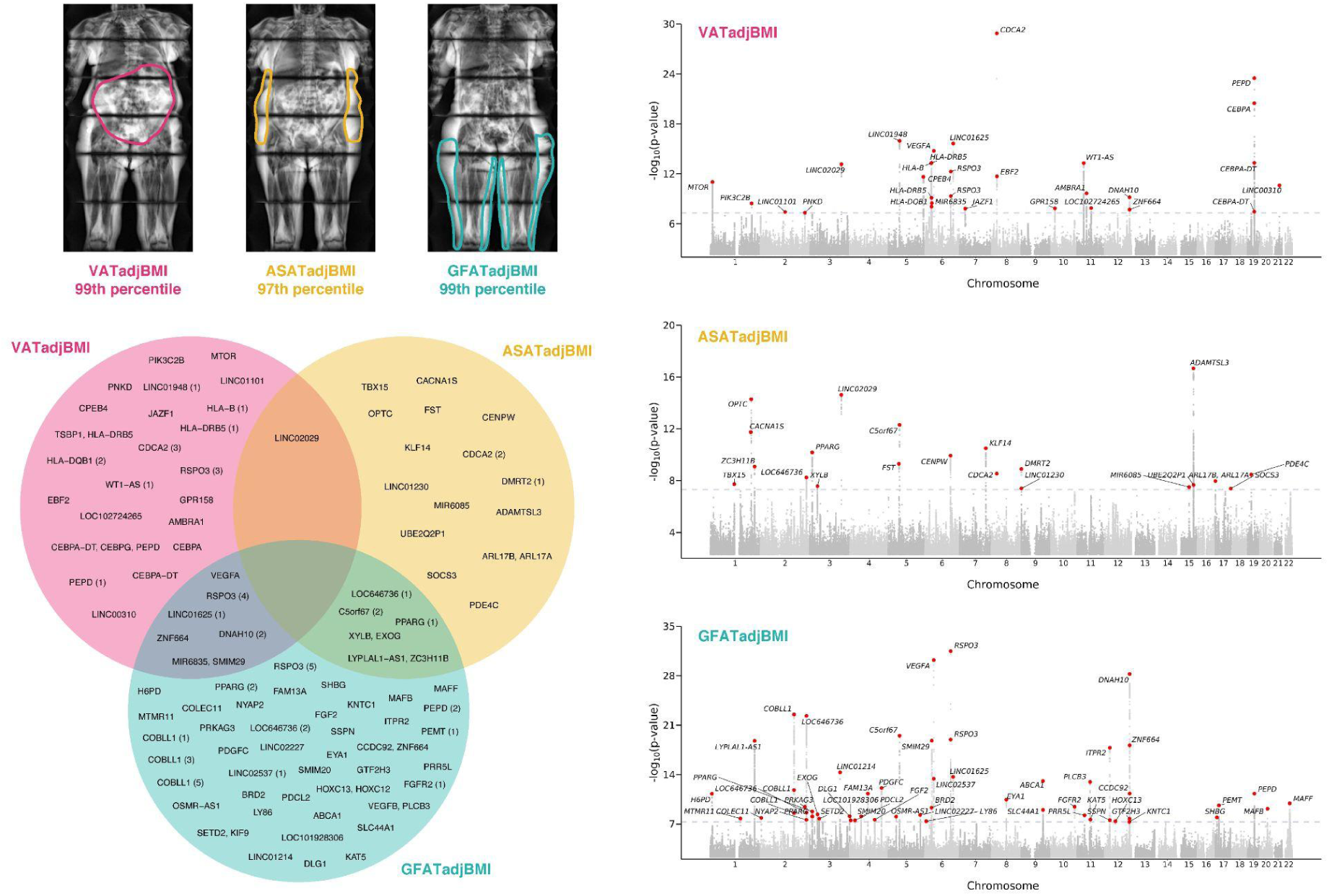
CVAS results for VATadjBMI, ASATadjBMI, and GFATadjBMI. (top left) Three females from the UK Biobank with similar age (67-70 years) and similar overweight BMI (27.6-28.6 kg/m^2^) with highly discordant fat distributions (right) Manhattan plots for sex-combined CVASs with VATadjBMI, ASATadjBMI, and GFATadjBMI. Lead SNPs are described in **Supplementary Table S4**. (bottom left) Overlap between VATadjBMI, ASATadjBMI, and GFATadjBMI loci denoted by the nearest gene (**Supplementary Table S5-6**).

The strongest association with ASATadjBMI was an intronic *ADAMTSL3* variant (rs768397327; P = 2.2 x 10^-17^), which was in near-perfect linkage disequilibrium (R^2^ = 0.97) with another intronic *ADAMTSL3* variant (rs11856122) previously associated with bioelectrical impedance-derived arm fat ratio, leg fat ratio, and trunk fat ratio, suggesting that this may be a general subcutaneous adiposity signal (**Figure 1**; **Supplementary Figure S7**).^13^ Another genome-wide significant signal was observed with an intronic *PPARG* variant (rs527620413; P = 6.8 x 10^-11^). Rare variants in *PPARG* have previously been associated with familial partial lipodystrophy.^6, 7^ The minor alleles at this locus (MAF = 0.12), which additionally consisted of rs17036328 and rs71304101 (R^2^ > 0.90), led to a signature of increasing ASATadjBMI (beta = 0.071), increasing GFATadjBMI (beta = 0.062), decreasing VAT/ASAT ratio (beta = -0.08), and decreasing VAT/GFAT ratio (beta = -0.058). These data suggest that common variation at *PPARG* can lead to adiposity variation along the lipodystrophy axis – for this locus, the minor alleles associated with an “anti-lipodystrophic” phenotype. *FST* is another gene that promotes adipogenesis and may have a causal role in insulin resistance – an intronic variant in *FST* (rs55744247) associated with ASATadjBMI (P = 5.1 x 10^-10^), but not VATadjBMI (P = 0.80) or GFATadjBMI (P = 0.25).^41^ Finally, a newly-identified intronic *DMRT2* variant (rs6474550; P = 1.3 x 10^-9^) associated with ASATadjBMI. In a study investigating fat depot-specific transcriptome signatures before and after exercise, *DMRT2* was one of three genes with higher expression in ASAT versus GFAT both before and after exercise.^42^

The top GFATadjBMI signal was an intronic *RSPO3* variant (rs72959041; P = 3.2 x 10^-32^) that has previously been shown to be a top signal for WHRadjBMI (**Figure 1**; **Supplementary Figure S8**).^12^ Recent work clarified this SNP as the causal variant at the locus and suggested that the minor allele concurrently reduces leg fat mass and increases android fat mass^43^. Our results confirm and further clarify these findings – the minor allele (MAF = 0.05) led to marked reduction of GFATadjBMI (beta = - 0.195; P = 3.2 x 10^-32^) and increased of VATadjBMI (beta = 0.118; P = 7.8 x 10^-13^), but a nonsignificant effect on ASATadjBMI (beta = -0.029; P = 0.09). Three independent intronic *COBLL1* variants (R^2^ < 0.1) were associated with GFATadjBMI (rs13389219; P = 3.0 x 10^-23^, rs3820981; P = 1.5 x 10^-12^, rs34224594; P = 2.8 x 10^-9^), but not VATadjBMI (P_min_ = 0.009) or ASATadjBMI (P_min_ = 2.7 x 10^-3^). One of these variants (rs13389219) is in LD with another intronic *COBLL1* variant (rs6738627) which has previously been implicated in a metabolically healthy obesity phenotype characterized by increased HDL cholesterol and reduced triglycerides despite increased body fat percentage^44^. In this study, aligning rs13389219 to the BMI-increasing direction (beta = 0.011, P = 7.3 x 10^-3^) revealed a concurrent increase in GFATadjBMI (beta = 0.073), consistent with a metabolically healthy fat depot shift. Finally, a GFATadjBMI association was observed at an intronic *PDGFC* variant (rs6822892; P = 8.0 x 10^-13^) – *PDGFC* was recently prioritized as a candidate causal gene for insulin resistance in human preadipocytes and adipocytes^41^.

Several associations were exclusive to CVASs of fat depot ratios (**Supplementary Figures S9-11)**. A missense variant in *ACVR1C* significantly reduced VAT/GFAT ratio (rs55920843; beta = -0.18; P = 1.9 x 10^- 8^). Prior work demonstrated that sequence variation in *ACVR1C* – including this variant – reduces WHRadjBMI and risk of type 2 diabetes^45^. Another missense variant in *ACVR1C* was nominally associated with reduced VAT/GFAT ratio, strengthening the importance of this gene (rs56188432 (Ile195Thr); beta = -0.20 (95% CI: -0.36 - -0.05); P < 0.01) (**Supplementary Table S7**). Another association with reduced VAT/GFAT ratio was present with a missense variant in *SERPINA1* (rs28929474; beta = -0.16; P = 4.8 x 10^- 10^). This variant has previously been associated with increased ALT and cirrhosis^45^. A shift towards a metabolically healthy fat distribution alongside increased risk of cirrhosis is consistent with *SERPINA1*- variation mediated cirrhosis occurring in association with alpha-1-antitrypsin deficiency, rather than non-alcoholic fatty liver disease^46^.

### Sex heterogeneity in genetic associations wtih local adiposity traits

Given prior work has noted significant sex heterogeneity in the genetic basis of anthropometric traits, we next tested for such heterogeneity for each of the six local adiposity traits.^11, 12, 47, 48^ Genetic correlations between sex-stratified summary statistics indicated overall high correlation between traits, with r_g_ somewhat higher for VATadjBMI (r_g_ = 0.87) as compared to ASATadjBMI or GFATadjBMI (r_g_ = 0.80 and 0.79 respectively) (**Supplementary Table S8**). We next tested for sex-dimorphism across individual loci that were genome-wide significant for either sex-combined or sex-stratified analyses for each local adiposity trait (**Figure 2**, **Supplementary Figure S12, Supplementary Table S9**). Three of 39 VATadjBMI loci (8%), six of 27 ASATadjBMI loci (22%), and nine of 66 GFATadjBMI (14%) showed significant sex dimorphism (P_diff_ < 0.05/158 = 3.2 x 10^-4^). The majority of these signals were driven by a greater magnitude of effect in females, which is consistent with prior investigations of WHRadjBMI^12, 48^. Across all six local adiposity traits, 25 loci were only genome-wide significant in females, while 9 loci were only genome-wide significant in males.

**FIGURE 2.**
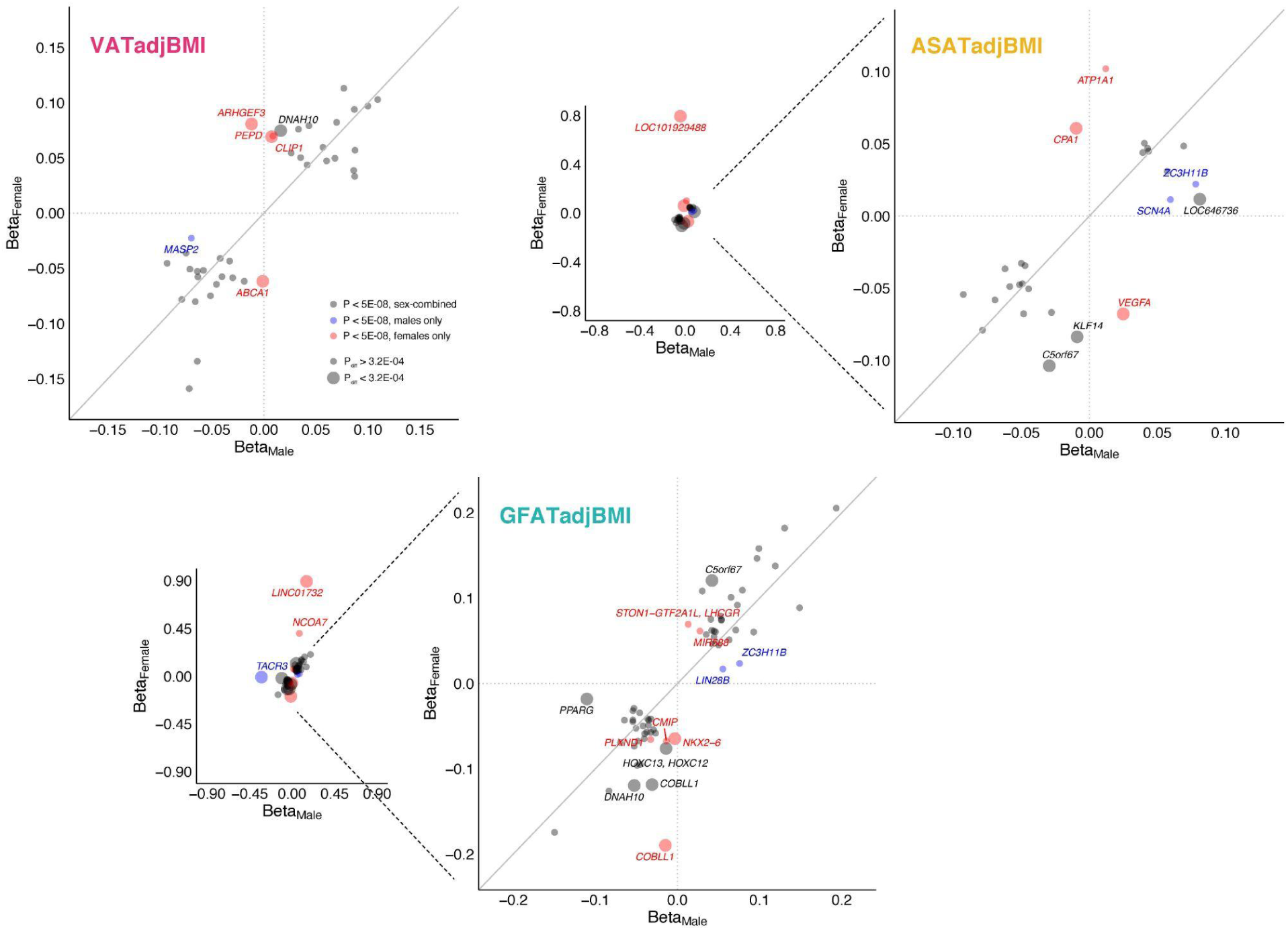
Common variant sex heterogeneity for VATadjBMI, ASATadjBMI, and GFATadjBMI local adiposity traits. For each adiposity trait, loci that were associated with the trait in either sex-combined or sex-stratified analyses are plotted (**Supplementary Table S7**). 39 such loci are plotted for VATadjBMI, 27 for ASATadjBMI, and 66 for GFATadjBMI. Loci colored black were genome-wide significant (P < 5 x 10^-8^) in sex-combined analysis, blue loci were significant for males, but neither females nor sex-combined, and red loci were significant for females, but neither males nor sex-combined. P_diff_ indicates the P-value for a hypothesis test comparing SNP effects in males and females, as implemented in EasyStrata software (Methods). 158 unique loci were tested for sex heterogeneity across the three adjusted-for-BMI traits and three fat depot ratios (**Supplementary Figure S12**), so a significance threshold of P_diff_ < 0.05/158 = 3.2 x 10^-4^ was set – large circles indicate that a given locus met this criterion.

### Overlap of local adiposity traits with WHRadjBMI findings

To investigate the added value of precisely quantifying fat depots with MRI in a smaller number of individuals as compared to WHRadjBMI in a larger cohort, we studied the effects of 345 loci identified in the most recent WHRadjBMI meta-analysis of up to 694,649 individuals on VATadjBMI, ASATadjBMI, and GFATadjBMI (**Figure 3, Supplementary Table S10**)^12^. Of the 345 loci, 10 (3%) achieved genome-wide significance in association with VATadjBMI (P < 5 x 10^-8^), 2 with ASATadjBMI (0.6%), and 14 (4%) with GFATadjBMI. A unit increase in WHRadjBMI might be expected to be reflecting a unit increase in VATadjBMI or ASATadjBMI, or a unit decrease in GFATadjBMI. We quantified how often a locus was discordant from this pattern (e.g. a unit increase in WHRadjBMI corresponding to a unit decrease in VATadjBMI), excluding loci where the fat depot effect size was smaller in magnitude than the standard error. Fifteen of 242 loci (6%) were VATadjBMI-discordant, 71 of 166 loci (43%) were ASATadjBMI-discordant, and 22 of 231 loci (10%) were GFATadjBMI-discordant.

**FIGURE 3.**
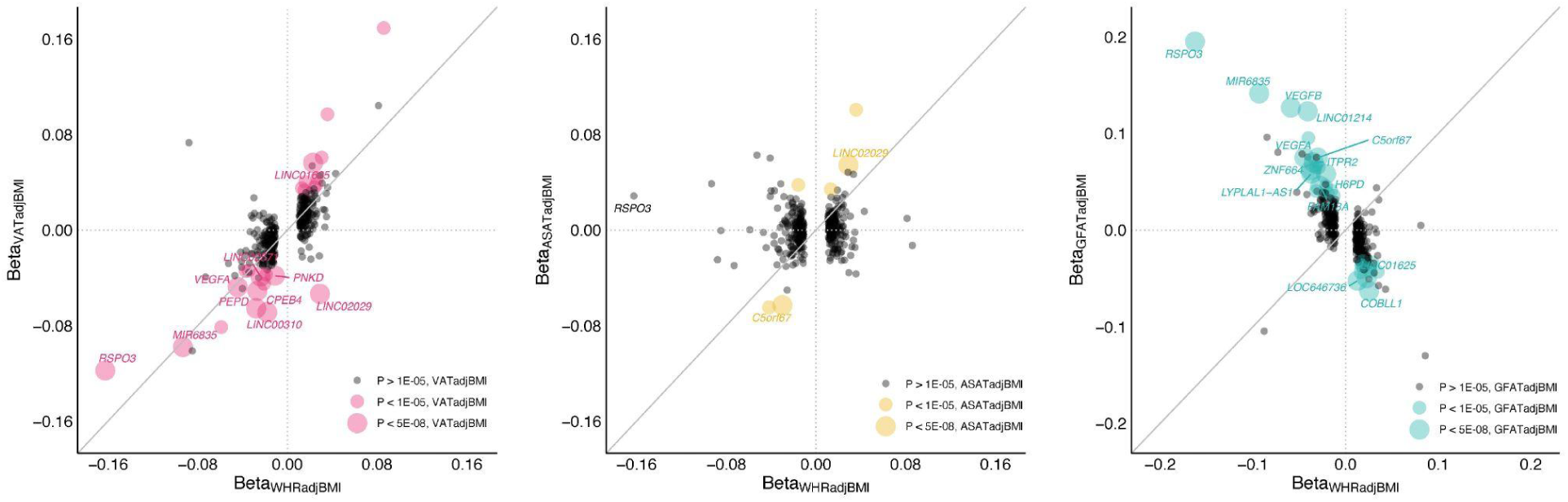
Effects of previously identified WHRadjBMI loci on local adiposity traits. 346 index SNPs associated with WHRadjBMI in a recent meta-analysis from the GIANT consortium were studied here -- one of these, rs139271800 (A>G) was not available in the studied cohort.^12^ Of the remaining 345 SNPs, effect sizes of VATadjBMI, ASATadjBMI, and GFATadjBMI are plotted against the effect size for WHRadjBMI as reported in the cited study (**Supplementary Table S10**).

Two illustrative examples indicate how follow-up of WHRadjBMI associations from a very large study in a smaller study with local fat depots quantified may prove useful. The strongest WHRadjBMI signal is located at an intronic *RSPO3* locus (rs72959041; beta = -0.162; P = 2.1 x 10^-293^) – our work further clarifies that this signal is driven by an effect on VATadjBMI (beta = -0.118; P = 7.8 x 10^-13^) and GFATadjBMI (beta = 0.195; P = 3.2 x 10^-32^), but not ASATadjBMI (beta = -0.029; P = 0.09). In contrast, a WHRadjBMI signal located at a variant upstream of *LINC02029* (rs10049088; beta = 0.029; P = 1.5 x 10^-59^) is driven by ASATadjBMI (beta = 0.054; P = 7.3 x 10^-14^) and GFATadjBMI (beta = -0.034, P = 6.0 x 10^-6^), but has a VATadjBMI-discordant signal (beta = -0.053, P = 8.7 x 10^-13^).

### Transcriptome-wide association study

To prioritize genes, we conducted a transcriptome-wide association study (TWAS) based on gene expression data from visceral adipose tissue and subcutaneous adipose tissue from GTEx v7.^49^ Across all traits, the strongest TWAS association was observed with GFATadjBMI at an intronic *DNAH10* variant (rs7133378; GFATadjBMI P = 5.6 x 10^-29^), corresponding to expression of *CCDC92* (TWAS Z-score = 12.0; TWAS P = 2.7 x 10^-33^), *DNAH10OS* (Z-score = 10.5; P = 8.2 x 10^-26^), *RP11-380L11.4* (Z-score = 10.0; P = 2.0 x 10^-23^), *ZNF664* (Z-score = 8.8; 1.5 x 10^-18^), and *DNAH10* (Z-score = 7.9; 3.5 x 10^-15^) in subcutaneous adipose tissue (**Supplementary Table S11**). In an adipocyte model, knockdown of *CCDC92* or *DNAH10* led to significant reduction of lipid accumulation, consistent with the direction of the TWAS Z-scores at this locus^19^. Of note, TWAS Z-scores at this locus were negative for VATadjBMI in visceral adipose tissue (*CCDC92* Z-score = -6.7; P = 2.7 x 10^-11^).

Another top TWAS signal was observed with GFATadjBMI at rs2713552, corresponding to expression of *IRS1* (Z-score = 9.1; P = 6.2 x 10^-20^). Prior work has demonstrated that decreased *IRS1* expression causes insulin resistance – our work further suggests that impaired expansion of the gluteofemoral depot may be involved in this physiological insult^41, 50^.

### Cell-specific enrichment analyses

We used stratified LD-score regression to probe for cell- and tissue-specific enrichment for each adiposity trait (**Supplementary Table S12**).^51^ A marked dichotomy was observed between the three global adiposity traits (VAT, ASAT, GFAT) and the six local adiposity traits (VATadjBMI, ASATadjBMI, GFATadjBMI, VAT/ASAT, VAT/GFAT, ASAT/GFAT). While VAT, ASAT, and GFAT showed a pattern of central nervous system (CNS) tissue enrichment – consistent with the enrichment pattern for BMI – local adiposity traits were characterized by adipose tissue signals with reduced CNS signals (**Supplementary Figures S13-14**). These results further emphasize that the genetic basis of overall weight and adiposity is driven largely by central nervous system processes – such as those governing appetite and satiety – whereas fat distribution is regulated at the level of the adipocyte and other peripheral tissues.

### Rare variant association study

Up to 19,255 individuals with fat depots quantified and exome sequencing data available were included in rare variant association studies. We utilized two masks: one containing only predicted loss-of-function variants (pLoF) and a second combining pLoF with missense variants predicted to be deleterious by 5 out of 5 *in silico* prediction algorithms (pLoF+missense). We tested the association between the aggregated rare variant score with each mask and each inverse normal transformed phenotype using multivariable regression. Analyses were restricted to genes with at least 10 variant carriers in the analyzed cohort, yielding 12,020 tested genes and an exome-wide significance threshold of P < 0.05/12,020 = 4.2 x 10^-6^. One exome-wide significant association was identified: pLoF+missense variants in *PDE3B* associated with increased GFATadjBMI in females (24 carriers; beta = 0.98; P = 1.7 x 10^-6^) (**Supplementary Table S13**). Individuals who carry loss-of-function variants in *PDE3B* have previously been demonstrated to have reduced WHRadjBMI^52^. This study confirms and extends this result by demonstrating that females, and not males, who carry pLoF+missense variants in *PDE3B* demonstrate increased GFATadjBMI, reduced VATadjBMI, and a nonsignificant change in ASATadjBMI, indicating that inherited deficiency of this gene is associated with a metabolically favorable fat distribution (**Figure 4**; **Supplementary Table S14**).

**FIGURE 4.**
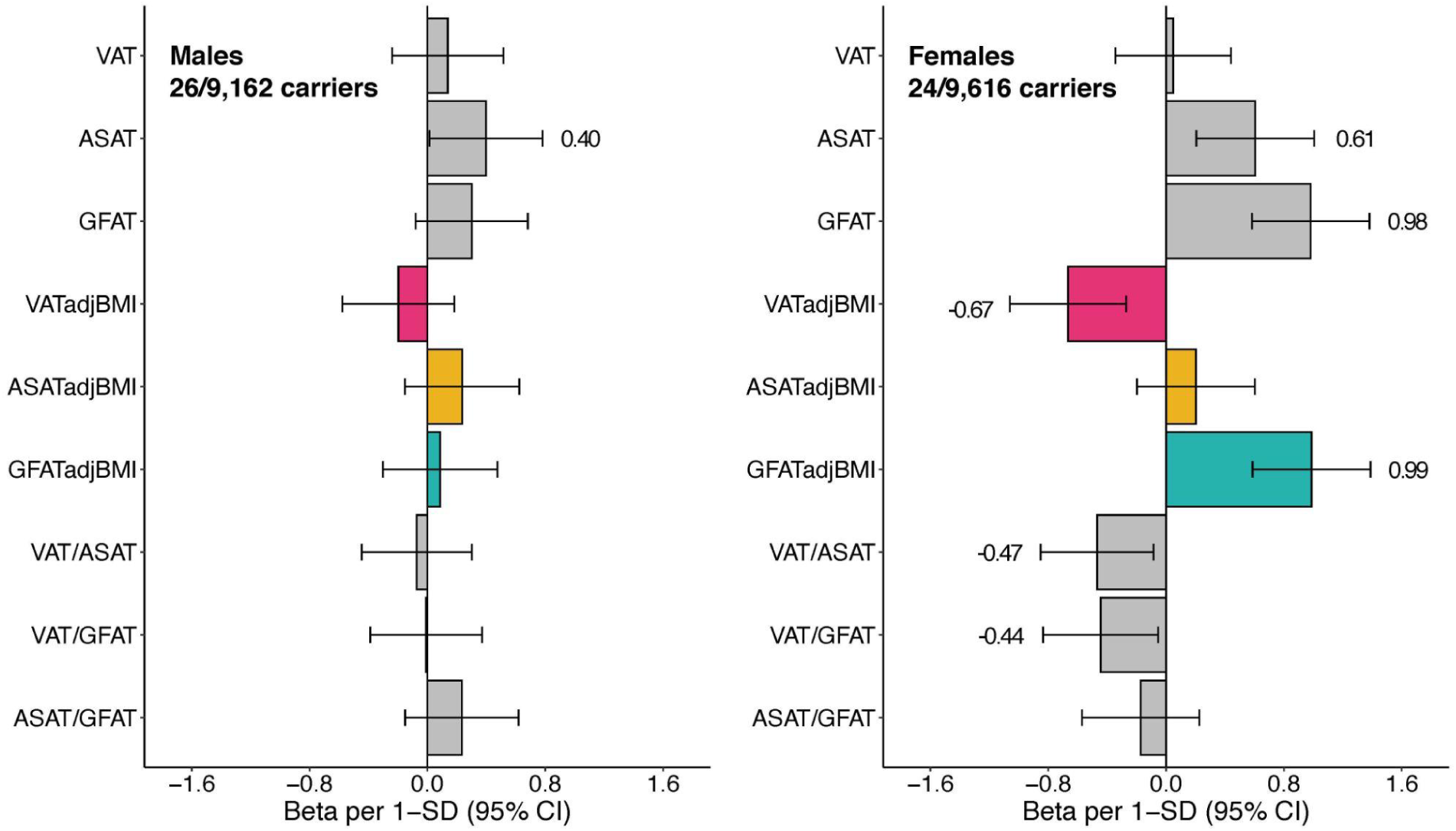
Predicted loss-of-function and missense rare variants in PDE3B selectively modulate fat distribution in females. A mask combining predicted loss-of-function variants and missense variants in *PDE3B* associated with GFATadjBMI in females with exome-wide significance (**Supplementary Table S13**). Carriers of these variants were separately associated with each adiposity phenotype in males and females in linear regressions adjusted for age, age squared, imaging center, genotyping array, and the first 10 principal components of genetic ancestry - the results are reported in **Supplementary Table S14** and plotted here. Associations that are nominally significant (p < 0.05) have the corresponding effect size written adjacent to the bar.

Rare variant signals in two additional genes, while they did not reach our threshold for statistical significance, warrant discussion. pLoF+missense variants in *ACAT1* associated with VAT in females (23 carriers; beta = 2.66; P = 6.4 x 10^-6^) and pLoF+missense variants in *PCSK1* associated with GFAT in sex-combined analysis (101 carriers; beta = 1.11; P = 7.5 x 10^-6^). Both of these genes have previously been implicated in altering adiposity. Rare mutations in *PCSK1* are known to cause monogenic obesity.^53, 54^ In a study comparing obese women with or without type 2 diabetes, gene expression of *ACAT1* was downregulated in the VAT and ASAT of obese women with type 2 diabetes and expression was restored after bariatric surgery and weight loss, suggesting a role in obesity-associated insulin resistance^55^.

### Polygenic contribution to extremes of fat distribution traits

Because many individuals with lipodystrophy-like phenotypes – especially in it more subtle forms – do not harbor a known pathogenic rare variant, prior studies have started to explore a potential “polygenic lipodystrophy,” in which inherited component is instead driven by the cumulative impact of many common DNA variants.^10, 19, 20, 56^ We set out to further test this hypothesis by generating polygenic scores consisting of up to 1,125,301 variants for VATadjBMI, ASATadjBMI, and GFATadjBMI traits using the LDPred2 algorithm.^57^ To ensure no overlap between summary statistics and tested individuals, CVAS was conducted using a randomly selected 70% of participants. An additional 10% of participants was used as training data to select optimal LDPred2 hyperparameters and the remaining 20% of participants were held out for testing. Participants at the tails of the distribution for any of the three local adiposity metrics were enriched in extreme polygenic scores – for example, participants in the top 5% of the GFATadjBMI distribution were nearly four times as likely to have a GFATadjBMI polygenic score in the top 5% of the distribution (OR = 3.81; 95% CI: 2.76-5.17) (**Figure 5**). Conversely, individuals with less than the 5th percentile of GFATadjBMI were over three times as likely to have a GFATadjBMI polygenic score less than the 5th percentile (OR = 3.36; 95% CI: 2.32-4.77). These findings suggest that polygenic inheritance plays an important role in fat distribution, with effect size particularly pronounced among those with more extreme imaging phenotypes.

**FIGURE 5.**
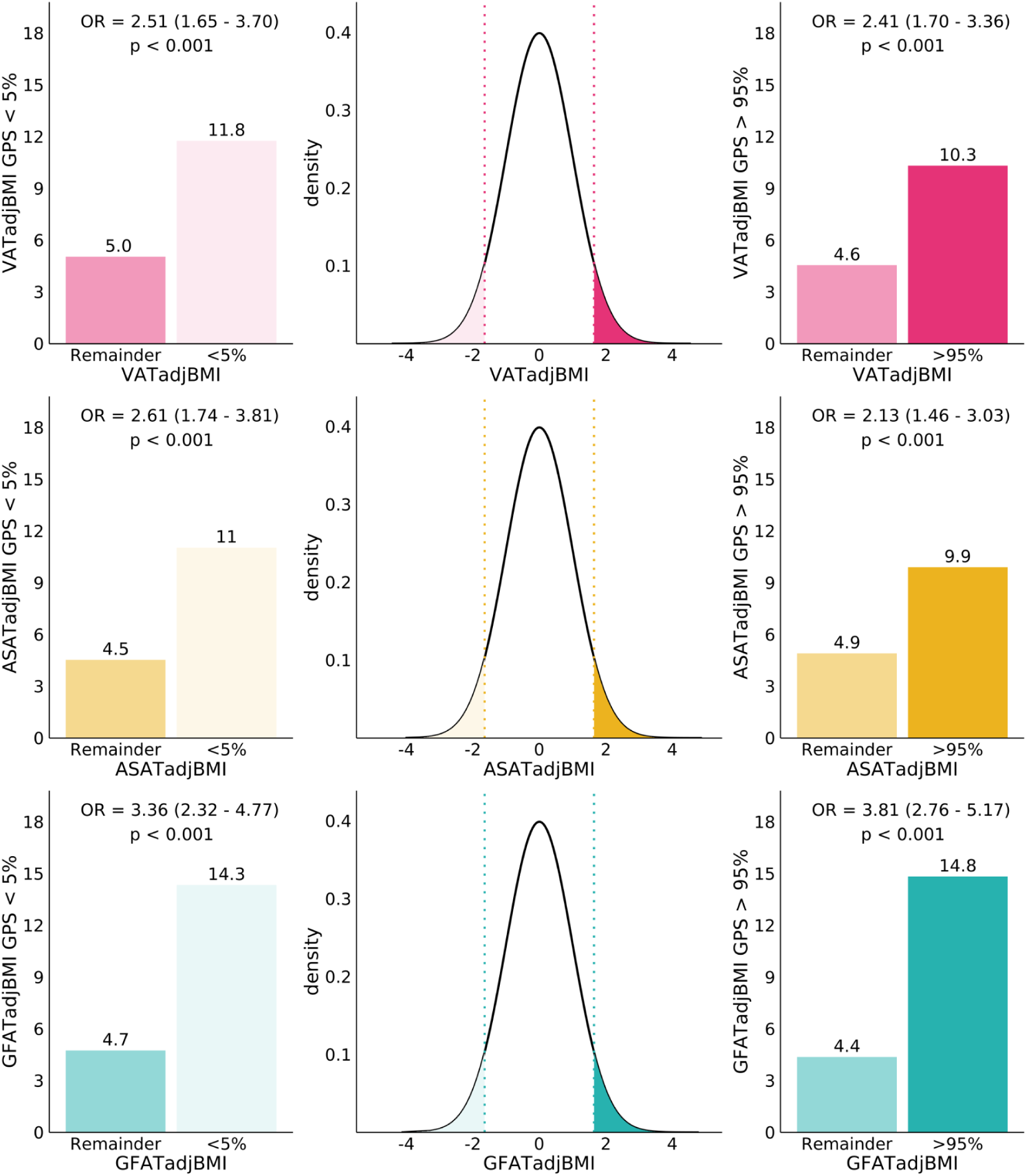
Enrichment of VATadjBMI, ASATadjBMI, and GFATadjBMI genome-wide polygenic scores in tails of the distribution. For each fat depot adjBMI trait, a polygenic score was trained using LDpred2 on 70% of the studied cohort and a 10% validation cohort was used to determine the optimal set of hyperparameters. Results in this figure and Figure 6 correspond to the 20% holdout cohort (N = 7,795).

We next tested the relationship between local adiposity polygenic scores and biomarkers of metabolic health (hemoglobin A1C, HDL cholesterol, serum triglycerides, and alanine aminotransferase (ALT)) and disease outcomes (type 2 diabetes, hypertension, and coronary artery disease) (**Figure 6**; **Supplementary Table S15**). Within the held out testing dataset, individuals in the top 10% of the GFATadjBMI polygenic score had lower hemoglobin A1C (beta: -0.10; 95% CI: -0.17 – 0.02; P = 9.9 x 10^-3^), higher HDL-cholesterol (beta: 0.14; 95% CI: 0.06-0.22; P = 4.3 x 10^-4^), lower serum triglycerides (beta: - 0.16; 95% CI: -0.24--0.09; P = 2.4 x 10^-5^), lower serum ALT (beta: -0.11; 95% CI: -0.19 – 0.04; P = 0.004), lower risk of type 2 diabetes (OR: 0.60; 95% CI: 0.39 – 0.90; P = 0.01), and lower risk of hypertension (OR: 0.81; 95% CI: 0.68 – 0.96; P = 0.02). By contrast, those in the top 10% of the VATadjBMI polygenic score tended to have increased risk of these disease outcomes with odds ratios for type 2 diabetes, coronary artery disease, and hypertension of 1.62, 1.10, and 1.07 respectively.

**FIGURE 6.**
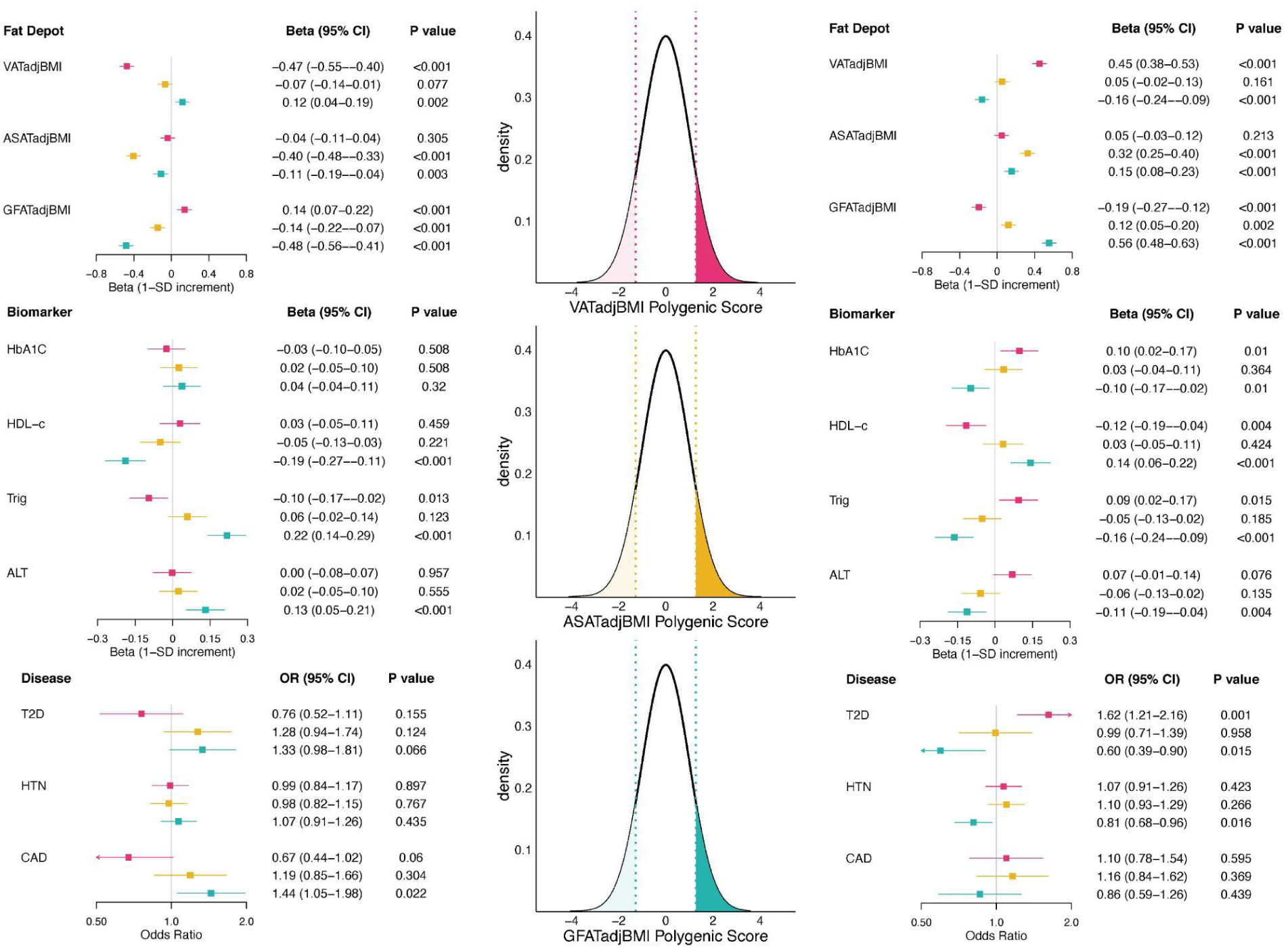
Effects of VATadjBMI, ASATadjBMI, and GFATadjBMI polygenic scores on metabolically relevant biomarkers and diseases. The central density plots indicate the distributions of VATadjBMI, ASATadjBMI, and GFATadjBMI polygenic scores in a subset of the imaged and genotyped individuals in this study who were held out during polygenic score development (20% of the cohort, N = 7,795). The dotted lines and shaded regions correspond to individuals in the top 10% and bottom 10% of the polygenic score. Forest plots to the right correspond to effect sizes of an indicator variable for being in the top 10% of the polygenic score (with identical color-coding to the density plots), while forest plots to the left correspond to effect sizes of an indicator variable for being in the bottom 10% of the polygenic score. Each polygenic score was residualized against the first 10 principal components of genetic ancestry prior to regression, and each regression was adjusted for age at imaging, sex, and the first 10 principal components of genetic ancestry. HbA1C, hemoglobin A1C; HDL-c, HDL-cholesterol; Trig, triglycerides; ALT, alanine aminotransferase; T2D, prevalent type 2 diabetes (at time of imaging); HTN, prevalent hypertension; CAD, prevalent coronary artery disease. Corresponding data are found in **Supplementary Table S15.**

Among an independent dataset of 447,486 individuals of the UK Biobank who were genotyped, but not imaged, these polygenic scores had consistent trends across metabolically relevant biomarkers and diseases (**Supplementary Figure S15; Supplementary Table S16**).

## DISCUSSION

In this study, we studied the inherited basis of body fat distribution as measured using abdominal MRI in up to 38,965 individuals. Results support significant heritability of derived measures of local adiposity, with 178 unique loci identified. These loci frequently demonstrated differential impact in males versus female participants and provided more precise assessment of previously identified loci for anthropometric proxies such as WHRadjBMI. Polygenic scores for local adiposity traits were highly enriched among those with ‘lipodystrophy-like’ fat distributions and were strongly associated with cardiometabolic traits in a depot-specific fashion. These results have at least three implications.

First, careful quantification of local adiposity measures using abdominal MRI and machine learning techniques enabled refinement of prior associations based on anthropometric proxies and discovery of new associations. Consistent with the raw fat depot volumes serving largely as proxies of overall adiposity, CVAS of VAT, ASAT, and GFAT each identified a well-known intronic *FTO* variant originally linked to BMI as a top signal. Cell-enrichment analyses corroborated these findings with each unadjusted fat depot displaying a pattern of central nervous system cell enrichment, consistent with the signal for BMI.^51^ By contrast, adjusted-for-BMI fat depots and fat depot ratios – local adiposity traits – were more heritable than global adiposity traits, revealed depot-specific genetic architecture, and displayed a pattern of adipose tissue cell-enrichment. These results lay the scientific foundation for functional genomics work to enhance understanding of key biologic pathways driving fat distribution, as might be enabled by large scale perturbational assays of adipocyte cell lines derived from specific depots.

Second, gluteofemoral fat volume is highly heritable particularly in females, with a genetic architecture that is distinct from the visceral and abdominal subcutaneous fat depots. Common variant association studies of MRI-derived adiposity traits to date have been limited to the visceral and abdominal subcutaneous fat depots – in this study, only 11 of 54 genome-wide significant associations with GFATadjBMI were also associated with either VATadjBMI or ASATadjBMI.^23–25^ Owing to the high heritability of GFATadjBMI, a polygenic score based on this depot was a better predictor of GFATadjBMI than either of the polygenic scores for VATadjBMI or ASATadjBMI for their respective fat depots. Finally, a low GFATadjBMI polygenic score predicted a poor cardiometabolic profile, providing further evidence for the hypothesis that a primary insult in a metabolically unhealthy fat distribution is the inability of the gluteofemoral fat depot to adequately expand.^4, 58^

Third, this study extends prior work suggesting the existence of polygenic lipodystrophy and lays the groundwork for identifying individuals at high risk of this phenotype using polygenic scores of local adiposity traits.^10, 19, 20, 56^ While several of the familial partial lipodystrophies (FPLD) are known to be caused by monogenic variation in genes like *LMNA* and *PPARG*, FPLD type 1 has not been linked to a single mutation, leading some to suggest that this disease may be polygenic in nature.^10^ Lotta et al. provided evidence for this by demonstrating that individuals with FPLD1 had a higher burden of a 53- SNP insulin resistance polygenic score compared to the general population.^19^ In this study, individuals who harbor lower than average GFATadjBMI or ASATadjBMI or higher than average VATadjBMI may be conceptualized as having a mild lipodystrophy-like phenotype. We demonstrate that individuals at the extremes of these local adiposity traits are enriched in extreme polygenic scores suggesting that polygenic scores may be helpful in identifying this subgroup of individuals for future focused investigations. For example, growth hormone releasing hormone analogs – such as tesamorelin – have previously been shown to lead to a selective reduction of VAT in patients with obesity or HIV-associated lipodystrophy.^59, 60^ Whether a local adiposity polygenic score – perhaps in combination with emerging imaging tools for identifying lipodystrophies – could identify a subset of individuals with obesity and polygenic lipodystrophy who may benefit from these fat redistribution agents in addition to traditional obesity therapy is an area for future investigation.^61^

In conclusion, we carried out genetic association studies of local adiposity traits, including those derived from the gluteofemoral fat depot, in a large cohort of individuals with MRI imaging. Our work discusses the genetic architecture of the highly heritable gluteofemoral fat depot for the first time, and extends efforts to define and identify individuals with polygenic lipodystrophy.

## METHODS

### Study Population

The UK Biobank is an observational study that enrolled over 500,000 individuals between the ages of 40 and 69 years between 2006 and 2010, of whom 43,521 underwent MRI imaging between 2014 and 2020.^62, 63^ Our group previously estimated VAT, ASAT, and GFAT volumes in 40,032 individuals of the imaged cohort after excluding 3,489 (8.0%) scans based on technical problems or artifacts (**Supplementary Figure S1**).^5^ A subset of 39,076 individuals with genotype array data available was used as the primary cohort studied here. This analysis of data from the UK Biobank was approved by the Mass General Brigham institutional review board and was performed under UK Biobank application #7089.

### Deriving local adiposity traits

The focus of this study was to investigate the genetic architecture of fat distribution independent of the overall size of an individual. Two sets of traits were derived for this purpose: “adjBMI” traits and fat depot ratios. “adjBMI” traits represent residuals of the fat depot in question in sex-specific linear regressions against age, age squared, BMI, and height. We provide justification in the **Supplementary Methods** for adjusting for both BMI and height as opposed to only BMI. In brief, adjusting only for BMI introduces a significant genetic correlation of each adjBMI trait with height (most pronounced with ASAT and GFAT). Several prior studies have suggested that adjusting for heritable covariates can lead to spurious genetic associations due to collider bias.^64, 65^ We investigated the extent to which VATadjBMI, ASATadjBMI, and GFATadjBMI loci may be driven by collider bias with BMI or height and found little evidence for collider bias making a significant contribution to these results (**Supplementary Figures S16-18**; **Supplementary Table S17**).

### Genotyping, imputation, and QC

Genotyping in the UK Biobank was done with two custom genotyping arrays: UK BiLEVE and Axiom.^66^ Imputation was done using the UK10K and 1000 Genomes Phase 3 reference panels.^67, 68^ Prior to analysis, genotyped SNPs were filtered based on the following criteria, only including variants if: (1) MAF >= 1%, (2) Hardy-Weinberg equilibrium (HWE) P > 1 x 10^-15^, (3) genotyping rate >= 99%, and (4) LD pruning using R^2^ threshold of 0.9 with window size of 1000 markers and step size of 100 marker.^69,70^ This process resulted in 433,616 SNPs available for genetic relationship matrix (GRM) construction. Imputed SNPs with MAF < 0.005 or imputation quality (INFO) score < 0.3 were excluded. These criteria resulted in a total of 11,485,690 imputed variants available for analysis.

Participant were excluded from analysis if they met any of the following criteria: (1) mismatch between self-reported sex and sex chromosome count, (2) sex chromosome aneuploidy, (3) genotyping call rate < 0.95, or (4) were outliers for heterozygosity. Up to 38,965 participants were available for analysis (37,641 for adjBMI traits because these individuals also had to have BMI available).

### Common variant association studies (CVAS)

Nine traits were analyzed (VAT, ASAT, GFAT, VATadjBMI, ASATadjBMI, GFATadjBMI, VAT/ASAT, VAT/GFAT, and ASAT/GFAT) in three contexts (sex-combined, male only, female only), leading to 27 analyses in total. SNP-heritability was estimated using BOLT-LMM v2.3.4.^71, 72^ Genetic correlations between traits were estimated using cross-trait LD-score regression (*ldsc* v1.0.1) using default settings.^30, 31^

Prior to conducting genome-wide association studies, each trait was inverse-normal transformed. Each analysis was adjusted for age at the time of MRI, age squared, sex (except in sex-stratified analyses), the first 10 principal components of genetic ancestry, genotyping array, and MRI imaging center. BOLT-LMM v2.3.4 was used to carry out genome-wide association studies accounting for cryptic population structure and sample relatedness.^71, 72^ After the QC protocol detailed above, 433,616 SNPs were available for genetic relationship matrix (GRM) construction. A threshold of P < 5 x 10^-8^ was used to denote genome-wide significance, while a threshold of P < 5 x 10^-8^ / 27 = 1.9 x 10^-9^ was used to denote study-wide significance.

Lead SNPs were prioritized with LD clumping. LD clumping was done with the --clump function in PLINK to isolate independent signals for each CVAS. The parameters were as follows: --clump-p1 5E-08, --clump-p2 5E-06, --clump-r2 0.1, --clump-kb 1000, which can be interpreted as follows: variants with p < 5E-08 are chosen starting with the lowest p-value, and for each variant chosen, all other variants with p < 5E-06 within a 1000 kb region and r2 > 0.1 with the index variant are assigned to that index variant. This process is repeated until all variants with p < 5E-08 are assigned an LD clump. Note that in-sample LD was computed using a random sample of 3,000 individuals from the studied cohort to use as the LD reference panel for LD clumping.

The extent of genomic inflation versus polygenicity was assessed by computing the LD-score regression intercept (*ldsc* v1.0.1) using default settings.^30^

A lead SNP was defined as newly-identified if it was not in LD (R^2^ < 0.1) with any SNP in the CVAS catalog (downloaded June 08, 2021) with genome-wide significant association (P < 5 x 10^-8^) with any “DISEASE/TRAIT” containing the following characters: (1) “body mass”, (2) “BMI”, (3) “adipos”, (4) “fat”, (5) “waist”, (6) “hip circ”, or (7) “whr”. These characters captured key anthropometric traits of interest (e.g. body mass index, waist circumference, hip circumference, waist-to-hip ratio) as well as other related traits of interest (e.g. visceral adipose tissue, predicted visceral adipose tissue, fat impedance measures).

### Identification of sex-dimorphic signals

Genetic correlations between sexes for each of the adiposity traits were computed using cross-trait LD-score regression as described above.

Using sex-specific CVAS summary statistics for each of the six local adiposity traits (VATadjBMI, ASATadjBMI, GFATadjBMI, VAT/ASAT, VAT/GFAT, ASAT/GFAT), we tested each of the 158 genetic loci that were genome-wide significant for any of the six local adiposity traits in either sex-combined or sex-stratified analyses for sex dimorphism by computing the *t*-statistic:

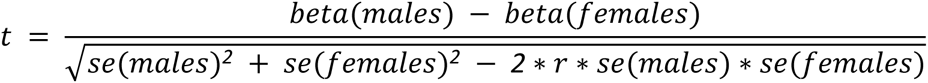

where beta is the effect size for an adiposity trait in sex-stratified CVAS, *se* is the standard error, and *r* is the genome-wide Spearman rank correlation coefficient between males and females. The *t*-statistic and associated P-value (P_diff_) were computed using the EasyStrata software.^73^ Given that 158 independent loci were tested, a significance threshold of P_diff_ < 0.05/158 = 3.2 x 10^-4^ was used.

### WHRadjBMI loci lookups

A recent meta-analysis for the WHRadjBMI trait across 694,649 individuals revealed 346 unique associated loci.^12^ Of these 346 loci, the primary signals for 345 loci were among the imputed variants available for analysis in this study. We plotted the effect sizes for VATadjBMI, ASATadjBMI, and GFATadjBMI for each of these 345 loci and further quantified the frequency of “WHRadjBMI- discordance” defined as either (1) WHRadjBMI and VATadjBMI effects going in opposite directions, (2) WHRadjBMI and ASATadjBMI effects going in opposite directions, or (3) WHRadjBMI and GFATadjBMI effects going in the same direction. For each adiposity trait in the “WHRadjBMI-discordance” analysis, we excluded loci for which the effect size beta was smaller than the standard error to avoid inflating the fraction of “WHRadjBMI-discordant” loci.

### Transcriptome-wide association study

For each of the six local adiposity traits (VATadjBMI, ASATadjBMI, GFATadjBMI, VAT/ASAT, VAT/GFAT, ASAT/GFAT), we performed a TWAS to prioritize genes on the basis of imputed cis-regulated gene expression using FUSION with default settings.^49, 74, 75^ Pre-computed gene expression weights from GTEx v7 were used as downloaded from the FUSION website (http://gusevlab.org/projects/fusion/).^49^ Reference weights for visceral adipose tissue were used for VATadjBMI, while those for subcutaneous adipose tissue were used for ASATadjBMI, GFATadjBMI, and ASAT/GFAT ratio. Weights from both visceral and subcutaneous adipose tissue were used for VAT/ASAT and VAT/GFAT ratios.

### Cell- and tissue-specific enrichment

We used stratified LD-score regression to identify cell types that are most relevant for each of the nine adiposity traits (VAT, ASAT, GFAT, VATadjBMI, ASATadjBMI, GFATadjBMI, VAT/ASAT, VAT/GFAT, and ASAT/GFAT) and BMI.^51^ We carried out this analysis using *ldsc* v1.0.1 with default settings and using two gene expression datasets that are described in the manuscript outlining stratified LD-score regression^51^: GTEx^76^ and the “Franke lab”^77, 78^ dataset.

### Sequencing and sample quality control for rare-variant association study

We conducted rare-variant association studies using data from the 200,643 exomes released by the UK Biobank.^79^ Whole-exome sequencing was performed by the Regeneron Genetics Center using an updated Functional Equivalence (FE) protocol that retains original quality scores in the CRAM files (referred to as the OQFE protocol) as previously described.^79^ The DTxGen Exome Research Panel v1.0 including supplemental probes was used for exome capture for this data set (https://biobank.ctsu.ox.ac.uk/showcase/label.cgi?id=170). 19,396 genes in the targets of 38Mbp were covered. 75×75bp paired-end reads were sequenced on the Illumina NovaSeq 6000 platform. For each sample in the targeted region, more than 95.2% of sites were covered by more than 20 reads. We downloaded the pVCF file provided by the UK Biobank, and then applied additional genotype call, variant, and sample quality control.

The individual genotype call was set as missing if reads depth (DP) ≤ 10 or DP ≥ 200, if homozygous reference allele with genotype quality (GQ) ≤ 20 or the ratio of alt allele reads over all of the covered reads > 0.1, if heterozygous with the ratio of alt allele reads over all of the covered reads < 0.2 or Phred-scaled likelihood (PL) of the reference allele < 20, or if homozygous alternate with the ratio of alt allele reads over all of the covered reads < 0.9 or PL of reference allele < 20. The variant quality control was performed using the following exclusion criteria:

- Variants in low-complexity regions of the genome that preclude accurate read alignment as previously defined.^80^
- Variants in segmental duplication region of the genome.^80, 81^
- Hardy-Weinberg disequilibrium (HWE) p-value < 1×10^-15^.
- Variant call rate < 90%.
- Monomorphic sites after the above genotype call quality control.

After the above genotype call and variant QC, we selected a subset of high-quality variants for inferring the genetic kinship matrix and genetic sex used for sample QC. We selected independent autosome variants by MAF > 0.1%, missingness < 1%, and HWE P > 10^-6^. We further pruned the variants using PLINK2 software^82^ with a window size of 200, step size 100, and R^2^ = 0.1 and removed indels and strand ambiguous SNPs. Based on these variants, we used KING (version 2.2.5)^83^ to infer the genetic kinship matrix. We further selected X-chromosomal variants, not within the pseudo-autosomal regions, based on the sample variant QC criteria as for the autosome variants and did the same variant pruning procedure. We then inferred the genetic sex based on the F statistics by PLINK2 software, F > 0.8 was set to male, while samples with F < 0.5 were set to female. 80 samples were removed because of the discordance of genetic sex with self-reported sex. We further removed samples if:

- The ratio of heterozygote/homozygote beyond 8 standard deviations (N = 100 samples removed).
- The ratio of the number of SNVs/indels beyond 8 standard deviations (N = 1 samples removed).
- The number of singletons was beyond 8 standard deviations (N = 111 samples removed).
- Genotype call rate < 90% (N = 1 sample removed)
- Withdrawal of informed consent (N = 13 samples removed)

We further randomly removed one sample if a pair of samples had second-degree relative or closer kinship, defined as kinship coefficient > 0.088474 (N = 1,563 samples removed). Of all the above QC passed samples, 19,255 samples out of the 40,032 having image-derived traits were used in the downstream rare variant burden test. We converted the genetic coordinates from GRCh38 to GRCh37 using CrossMap software (version: v0.3.3)^84^.

### Approach to variant annotation and weighting

To identify rare (minor allele frequency < 0.1%) high-confidence predicted inactivating variants, we applied the previously validated Loss-Of-Function Transcript Effect Estimator (LOFTEE) algorithm implemented within the Ensembl Variant Effect Predictor (VEP) software program as a plugin, VEP version 96.0^85, 86^. The LOFTEE algorithm identifies stop-gain, splice-site disrupting, and frameshift variants. The algorithm includes a series of flags for each variant class that collectively represent ‘low-confidence’ inactivating variants. In this study, we studied only variants that were ‘high-confidence’ inactivating variants without any flag values. This aggregation strategy will be referred to hereafter as putative loss-of-function (‘<u>pLoF</u>’).

To identify rare (minor allele frequency < 0.1%) predicted damaging missense variants, we included variants predicted to be damaging by all of five computational prediction algorithms as described previously^87–89^. In brief, predictions were retrieved from the dbNSFP database^90^, version 2.9.3, with the most severe prediction across multiple transcripts used. We focused on five prediction algorithms: SIFT^91^ (including variants annotated as damaging), PolyPhen2-HDIV and PolyPhen2-HVAR^92^ (including variants annotated as possibly or probably damaging), LRT^93^ (including variants annotated as deleterious), and MutationTaster^94^ (including variants annotated as disease-causing-automatic or disease-causing). Within the association testing framework, this class of variants was given a gene-specific weight based on the relative cumulative frequency of these predicted damaging missense variants as compared to the cumulative frequency of high-confidence predicted inactivating variants identified by LOFTEE algorithm using a previously recommended approach^95, 96^: given the cumulative allele frequency of all of the LOFTEE high confidence rare variants of a gene (*G*) as *f_L_*, the cumulative allele frequency of all of the predicted damaging missense variants as *f_M_*, the weight for the missense variants was estimated as 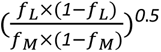, and capped at 1.0. For genes without LOFTEE high confidence rare variants, the weight for missense variants is 1.0. This aggregation strategy will be referred to hereafter as putative loss-of-function plus missense (‘<u>pLoF+missense</u>’).

### Statistical analysis

We tested the association between the aggregated rare variant score (the weighted sum of the qualified variant of each gene) and each inverse normal transformed phenotype using a multivariable regression model in sex-combined and sex-stratified models. Analyses were restricted to genes that had at least 10 variant carriers in the analyzed cohort. An individual’s gene-specific score was computed according to the weighting strategy described above and capped at one. The covariates were the same as the common variant association test. Given the filter of 10 variant carriers, sex-combined analyses tested 12,020 genes and so a gene was recognized as exome-wide significant if the gene’s *P-value* was smaller than the Bonferroni-corrected p-value threshold of 0.05/12,020 = 4.2 x 10^-6^.

### Polygenic score

We used the LDpred2 algorithm^57^ to derive genome-wide polygenic scores for each trait. We randomly selected 350,000 White British ancestry individuals from the UK Biobank to use as the LD reference panel,^66^ and used HapMap3 variants with MAF > 0.5% in the LD reference panel to compute the LD correlation matrix. For each trait, we partitioned the samples into three independent portions: 70% to run the CVAS for making the summary statistics, 10% to select the optimal hyperparameters, and 20% to test performance. We randomly removed one sample in a pair if the pair had a genetic relationship closer than a second-degree genetic relationship in the last two partitions of samples, and checked the pairwise relationship across the whole data set. For the hyperparameters of the LDpred2 algorithm, we grid searched three parameters: (1) 0.7, 1, and 1.4 times of genome-wide heritability estimation, (2) whether or not to use a sparse LD correlation matrix, and (3) 17 different estimates of the proportion of causal variants selecting from [0.18,0.32,0.56,1] x 10^[0,-1,-2,-3]^ and 0.0001. In total, we tested 3 x 2 x 17 = 102 grid points.

For all downstream analyses, each polygenic score was residualized against the first 10 principal components of genetic ancestry prior to regression with the dependent variable of interest, and each regression was adjusted for age at imaging, sex, and the first 10 principal components of genetic ancestry.

## Data Availability

The raw UK Biobank data is made available to researchers from universities and other research institutions with genuine research inquiries, following IRB and UK Biobank approval. Full GWAS summary statistics for each of the nine adiposity traits in sex-combined and sex-stratified analyses will be made available for download from the Broad Institute Cardiovascular Disease Knowledge Portal under the "Downloads" tab at http://www.broadcvdi.org/ at the time of publication.

## Author Contributions

S.A., M.W., and A.V.K. conceived and designed the study. S.A., M.W., M.D.R.K., J.S., and N.D. acquired, analyzed, and interpreted the data. S.A., M.W., and A.V.K. drafted the manuscript. S.A., M.W., M.D.R.K., J.S., H.D., N.D., S.H.C., S.J.J., P.T.E., A.P., K.N., M.C., P.B., and A.V.K. critically revised the manuscript for important intellectual content.

